# Comparative Effectiveness of Famotidine in Hospitalized COVID-19 Patients

**DOI:** 10.1101/2020.09.23.20199463

**Authors:** Azza Shoaibi, Stephen Fortin, Rachel Weinstein, Jesse A. Berlin, Patrick Ryan

## Abstract

**Background:** Famotidine has been posited as a potential treatment for COVID-19. We compared the incidence of COVID-19 outcomes (i.e., death; and death or intensive services use) among hospitalized famotidine users vs. proton pump inhibitors (PPIs) users, hydroxychloroquine users or famotidine non-users separately.

**Methods:** We constructed a retrospective cohort study using data from COVID-19 Premier Hospital electronic health records. Study population were COVID-19 hospitalized patients aged 18 years or older. Famotidine, PPI and hydroxychloroquine exposure groups were defined as patients dispensed any medication containing one of the three drugs on the day of admission. The famotidine non-user group was derived from the same source population with no history of exposure to any drug with famotidine as an active ingredient prior to or on the day of admission. Time-at-risk was defined based on the intention-to-treat principle starting 1 day after admission to 30 days after admission. For each study comparison group, we fit a propensity score (PS) model through large-scale regularized B logistic regression. The outcome was modeled using a survival model.

**Results:** We identified 2193 users of PPI, 5950 users of the hydroxychloroquine, 1816 users of famotidine and 26,820 non-famotidine users. After PS stratification, the hazard ratios for death were as follows: famotidine vs no famotidine HR 1.03 (0.89-1.18); vs PPIs: HR 1.14 (0.94-1.39); vs hydroxychloroquine:1.03 (0.85-1.24). Similar results were observed for the risk of death or intensive services use.

**Conclusion:** We found no evidence of a reduced risk of COVID-19 outcomes among hospitalized COVID-19 patients who used famotidine compared to those who did not or compared to PPI or hydroxychloroquine users.

## Introduction

Famotidine, a specific histamine-type 2 receptor antagonist that suppresses gastric acid production, has been proposed as an attractive candidate for COVID-19 treatment, based on the potential role that the histamine pathway may play in immune modulation^1^ and its long history of safe use^2^.

SARS-CoV-2 virus infection can induce histamine release via aberrant mast cell activation. The pathological histamine release has a potential to provoke the excessive synthesis of proinflammatory cytokines that may lead to acute respiratory distress syndrome observed in patients with severe COVID-19^1^. The finding of lower levels of inflammatory markers (e.g. C-reactive protein, ferritin, etc.) in famotidine-treated patients with COVID-19 suggests that the drug can dampen an uncontrolled proinflammatory immune response. This has prompted recent studies to postulate that the above effect of famotidine can be mediated via the antagonism and/or inverse agonism of histamine type 2 receptors ^3^. In another bioinformatics study, Wu et al^4^ identified famotidine as one of the drugs most likely to inhibit the 3-chymotrypsin-like protease (3CLpro), which processes proteins essential for viral replication^5^. Furthermore, famotidine has been shown in vitro to inhibit human immunodeficiency virus replication. Famotidine is currently being tested under an IND waiver for treating COVID-19 in a double blind randomized clinical trial at high intravenous doses (360 mg/day) in combination with either hydroxychloroquine or remdesivir (ClinicalTrials.gov Identifier: NCT04370262).

Recently, several observational studies have investigated the effect of famotidine on COVID-19 outcomes but have been limited to single-institutional explorations of small samples with varying statistical methods and inconsistent results. Freedberg et al^6^ reported that 84 patients receiving famotidine during a hospital stay, had a significantly reduced risk of death or intubation when compared to patients who did not^6^. Another study using electronic health data from Hartford hospital, included 83 patients receiving famotidine, and produced similar estimates^7^. Additionally, a sequential case series suggested benefits of famotidine treatment in an outpatient setting^8^. In contrast to these findings, a territory-wide study in Hong Kong found no significant association between severe COVID-19 disease and use of famotidine^9^.

Real world data can potentially provide critical and timely evidence on the effectiveness of famotidine on improving COVID-19 outcomes. However, the design and interpretation of an observational population-level effect estimation study might be challenging. The appropriateness in the selection of a comparator and adequacy of adjustment of baseline covariates can impact the risk of selection bias and confounding. Given the recency of this pandemic, assessments are further challenged by insufficient sample size, which introduces random error and also can limit the fidelity of any statistical adjustment performance.

Using real world data from a large national database on hospitalized patients with COVID-19, we aimed to estimate and compare the incidence of COVID-19 outcomes (i.e., death; and death or intensive services) among hospitalized famotidine users vs. proton pump inhibitors (PPIs) users, hydroxychloroquine users or famotidine non-users separately.

## Methods

We conducted a prevalent-user comparative retrospective cohort study measuring the association between famotidine use and severity of COVID-19 outcomes among patients hospitalized with COVID-19 in the United states. This study was approved under protocol (CCSDIH002924) https://github.com/ohdsi-studies/Covid19EstimationFamotidine/blob/master/Protocol/Covid19EstimationFamotidineProtocol.pdf

## Data Source

The study population was comprised of hospitalized patients with a diagnosis of COVID-19 available in the COVID-19 Premier Hospital Database (PHD). The PHD contains complete clinical coding, hospital cost, and patient billing data from approximately 700 hospitals throughout the United States. It captures from 20 to 25% of all inpatient admissions in the US. Premier collects de-identified data from participating hospitals in its health care alliance. The hospitals included are nationally representative based on bed size, geographic region, location (urban/rural) and teaching hospital status. The database contains medications administered during the hospitalization; laboratory, diagnostic, and therapeutic services; and primary and secondary diagnoses for each patient’s hospitalization. Identifier-linked enrollment files provide demographic and payor information. Detailed service-level information for each hospital day is recorded; this includes details on medication and devices received. All data were standardized to the Observational Health and Data Sciences and Informatics (OHDSI) Observational Medical Outcomes Partnership (OMOP) Common Data Model (CDM) version 5.3. The full description of the extract, transform and load of the data can be found here: https://github.com/OHDSI/ETLCDMBuilder/blob/master/man/PREMIER/Premier_ETL_CDM_V5_3.doc. PHD contains deidentified patient information, is Health Insurance Portability and Accountability Act (HIPAA) compliant, and is considered exempt from institutional review board (IRB) approval^10^.

### Study period and follow-up

The study period started from 02/01/2020 and ended at the latest available date for all data in 2020 (05/30/2020). Figure 1. illustrates the retrospective study design schematic. As illustrated in Figure 1, follow-up for each of the cohorts started at an index date defined by the first inpatient admission (day 0). Time-at-risk was defined based on the intention-to-treat principle starting 1 day after admission and continuing up until the first of: outcome of interest, loss to follow-up or 30 days after admission.

**Figure 1:**
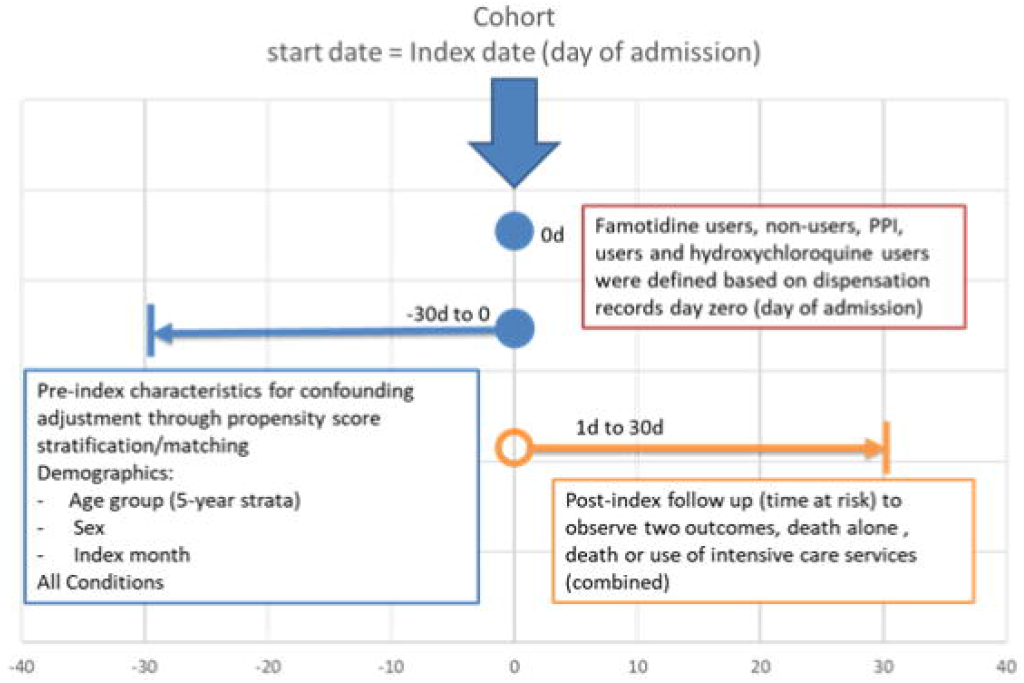
The study design schematic. We highlight index day specification, exposure definitions, adjustment strategies, outcome definitions and time-at-risk. Exposure involves prescriptions to drugs with RxNorm ingredients that map to famotidine, proton pump inhibitors (PPI) or hydroxychloroquine. The famotidine non-user group was derived from the same source population (participants) with no history of exposure to any drug with famotidine as an active ingredient prior to or on the day of admission.

### Study population

We included patients aged 18 years or older with an inpatient visit occurring after 02-01-2020; a condition, measurement or observation indicative of COVID-19 during or within 21 days prior to admission. Patients with evidence of intensive services (i.e., mechanical ventilation, tracheostomy or extracorporeal membrane oxygenation) at or within 30 days prior to admission were excluded.

### Exposures, outcomes and confounders

Famotidine, PPI and hydroxychloroquine exposure groups were defined as patients dispensed any medication containing one of the three drugs of interest -as an ingredient-on the day of admission. Patients who received both famotidine and any of the comparator drugs on day of admission were excluded. The famotidine non-user group was derived from the same source population (participants) with no history of exposure to any drug with famotidine as an active ingredient prior to or on the day of admission.

Outcomes of interest were death, identified based on patient discharge status within admission records, and death or intensive services (combined). Intensive services were defined as any condition, procedure or observation code indicative of mechanical ventilation, tracheostomy or extracorporeal membrane oxygenation. The code list used to identify study participants, exposures and outcomes can be found here https://github.com/ohdsi studies/Covid19EstimationFamotidine/blob/master/Protocol/Annex%20I%20-%20Concept%20Set%20Expressions.xlsx

## Statistical methods

To adjust for potential measured confounding and improve the balance between comparison cohorts, we built large-scale propensity score (PS) models for each comparison using regularized regression (Tian, Schuemie, and Suchard 2018). We used a Laplace prior (LASSO) with the optimal hyperparameter to fit the model, determined through 10-fold cross validation in which the outcome is a binary indicator for the potential comparator. This process used a large set of predefined baseline patient characteristics, including patient demographics (i.e., gender, age, index month) and all observed conditions within 30 days prior to or on admission. For computational efficiency, we excluded all features that occurred in fewer than 0.1% of patients within the target and comparator cohorts prior to PS model fitting. For the main analysis, we stratified into 5 PS strata and used conditional Cox proportional hazards models to estimate hazard ratios (HRs) between target and alternative comparator treatments for the risk of each outcome. The regression for the outcome models conditioned on the PS strata with treatment as the sole explanatory variable.

As a sensitivity analysis, we utilized a 1:1 PS matching and used an unconditional Cox proportional hazards model to estimate HRs in the matched set. We declared a HR as significantly different from no effect when its p < 0.05 without correcting for multiple testing.

Blinded to the results, the study team evaluated study diagnostics for these treatment comparisons to assess if they were likely to yield unbiased estimates. The suite of diagnostics included (1) minimum detectible risk ratio (MDRR), (2) preference score (a transformation of the PS that adjusts for prevalence differences between populations) distributions to evaluate empirical equipoise^11^ and population generalizability, (3) extensive patient characteristics to evaluate cohort balance before and after PS-adjustment. We defined target and comparator cohorts to achieve sufficient balance if all after-adjustment baseline characteristics return absolute standardized mean differences (SMD) < 0.1^12^.

We conducted this study using the open-source OHDSI CohortMethod R package (https://ohdsi.github.io/CohortMethod/) with large-scale analytics made possible through the Cyclops R package 13. The pre-specified protocol and start-to-finish open and executable source code are available at: https://github.com/ohdsi-studies/Covid19EstimationFamotidine. To promote transparency and facilitate sharing and exploration of the complete result set, an interactive web application https://data.ohdsi.org/Covid19EstimationFamotidine/ serves up study diagnostics and results for all study effects.

## Results

### Population and incidence

A total of 2193 users of PPI, 5950 users of the hydroxychloroquine, 1816 users of famotidine and 26,820 non-famotidine users were identified in the data and were eligible for the study. Table 1 illustrates patient cohort size, follow up duration, incidence of the two outcomes and MDRR for each famotidine/drug comparison for the PS stratified analysis. Among the famotidine group, a total of 1,331 (73.29%) and 374 (20.59%) patients received a dose of 20 and 40 mg of famotidine on the day of admission, respectively. Furthermore, 1,155 (63.60%) and 709 (39.04%) patients received oral and IV formulations of famotidine on the day of admission, respectively, with 1.4% (n=25) of patients receiving both oral and IV formulations.

**Table 1.**
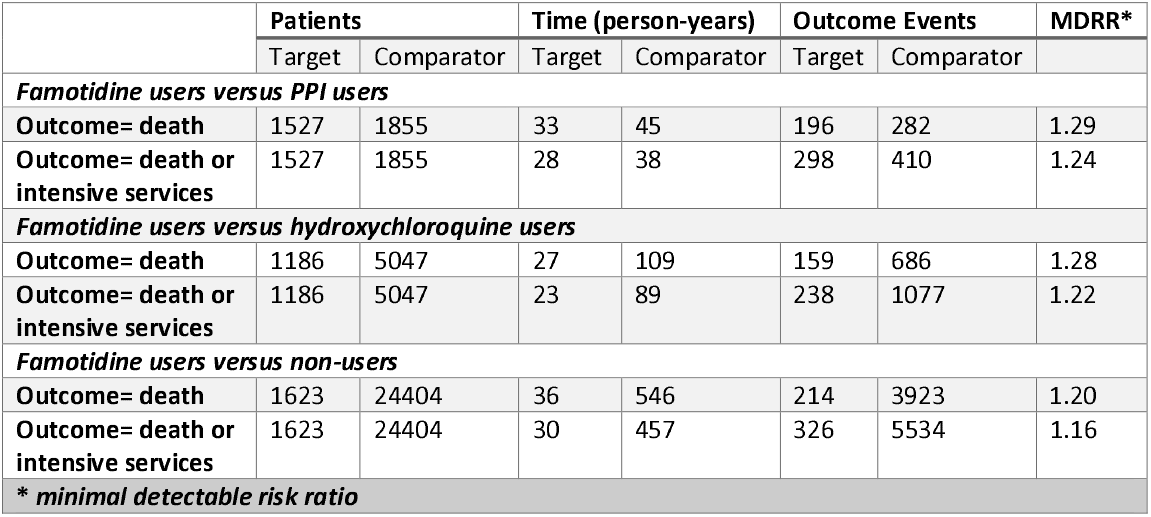
Populations and death events for proton pump inhibitors (PPI) users, hydroxychloroquine users, famotidine users and non-users, we report population size, total exposure time, outcome events and minimal detectable risk ratio (MDRR) for the PS stratified analysis.

### Famotidine users versus PPI users

After PS stratification, a total of 1,527 COVID-19 patients exposed to famotidine were compared to 1,855 patients exposed to PPI. Among famotidine users, the incidence of death alone was 12.83% (196 patients) vs 15.20% (282 patients) among PPI users. The incidence of death or intensive services (combined) was 18.96% (298 patients) vs 22.10% (410 patients) among famotidine and PPI users respectively.

### Famotidine users versus hydroxychloroquine users

After PS stratification, A total of 1,186 COVID-19 patients exposed to famotidine were compared to 5,047 patients exposed to hydroxychloroquine. Among famotidine users, the incidence of death alone was 13.40% (159 patients) vs 13.59% (686 patients) among hydroxychloroquine users. The incidence of death or intensive services (combined) was 20.07% (238 patients) vs 22.10% (1077 patients) among famotidine and hydroxychloroquine users respectively.

### Famotidine users versus non-users

After PS stratification, A total of 1,623 COVID-19 patients exposed to famotidine were compared to 24,404 patients in the famotidine non-user group. Among famotidine users, the incidence of death alone was 13.19% (214 patients) vs 16.09% (3923 patients) among non-users. The incidence of death or intensive services (combined) was 20.09% (326 patients) vs 22.68% (5534 patients) among famotidine users and non-users respectively.

### Characteristics of patients

Selected baseline characteristics of famotidine users compared to PPI users before and after PS stratification are shown in Table 2. Compared to PPI users, prior to propensity score adjustment, famotidine users were younger and had fewer comorbid conditions (based on (SMD) > 0.1). Specifically, PPI users were more likely to have chronic liver diseases, chronic obstructive lung disease, gastroesophageal reflux disease (GERD), gastrointestinal hemorrhage, hyperlipidemia, lesion of liver, osteoarthritis, renal impairment, rheumatoid arthritis, viral hepatitis C and cardiovascular disease.

**Table 2.**
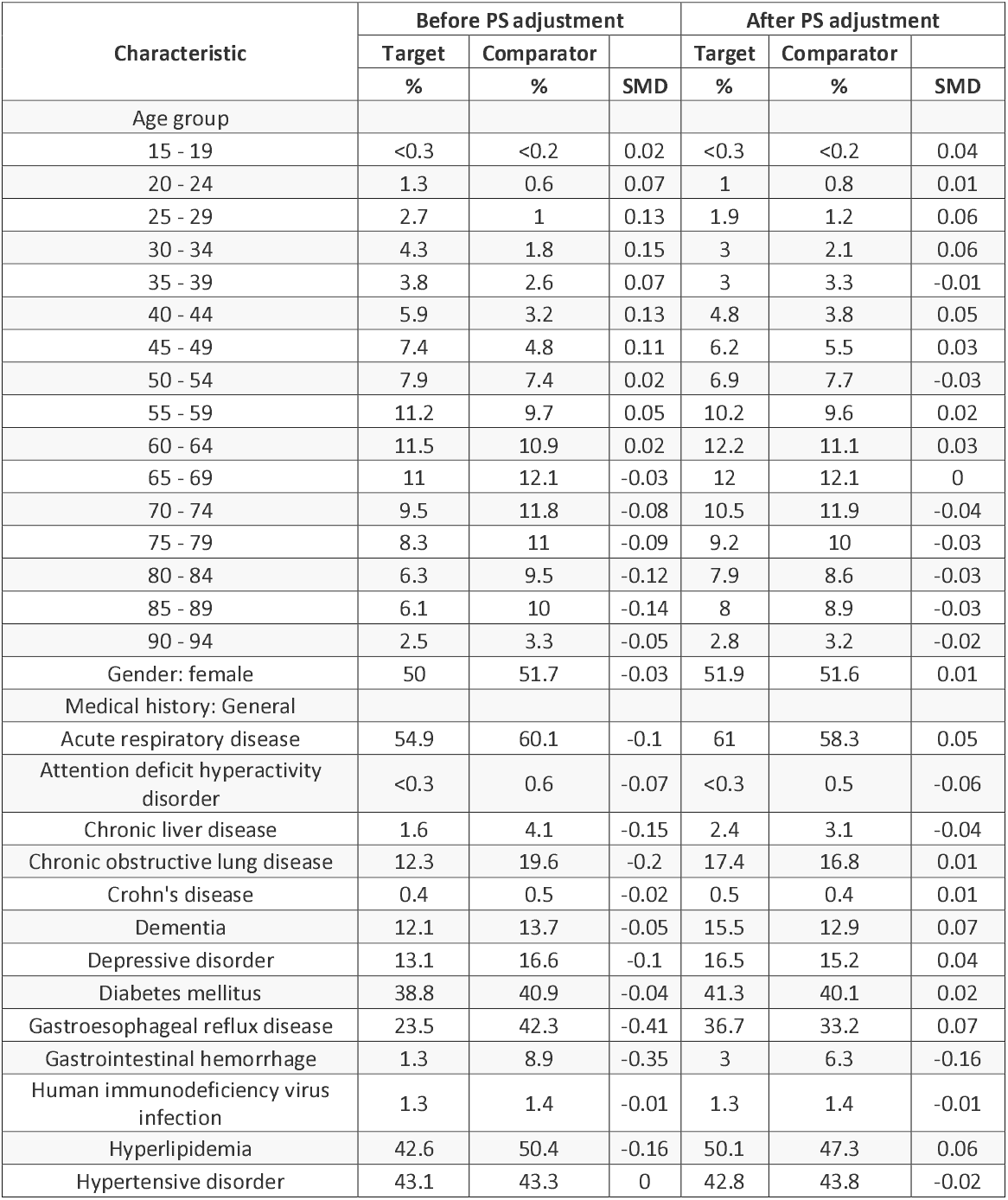
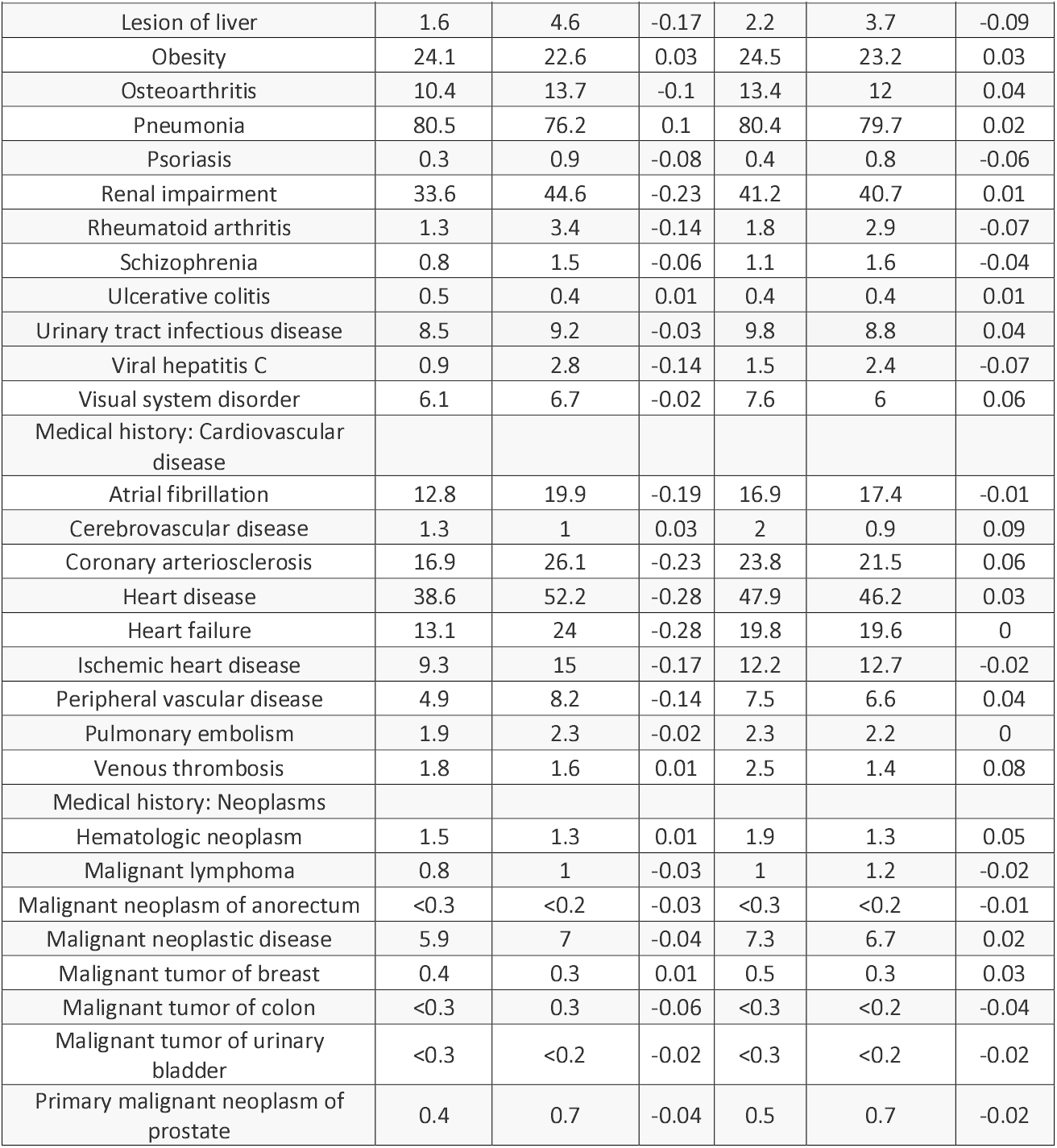
Cohorts characteristics of famotidine and proton pump inhibitor (PPI) users. We report the proportion of based-line characteristics and the standardized mean difference (SMD) before and after stratification. Less extreme SMD through stratification suggest improved balance between patient cohorts through propensity score adjustment

Selected baseline characteristics of famotidine users compared hydroxychloroquine users and to famotidine non-users, before and after PS stratification, are shown in Supplement Tables 1 and 2 respectively. Compared to hydroxychloroquine users, prior to propensity score adjustment, famotidine users were more likely to be females, have GERD, some cardiovascular diseases, urinary tract infections and depressive disorder but less likely to have pneumonia. Compared to non-users, famotidine users were less likely to be in the older age group (85-89 years) but were more likely to have GERD.

### PS model adjustment and cohort balance

More than 2400 baseline patients’ characteristics were available for PS adjustment. After large-scale PS stratification or matching, SMDs for most baseline characteristics were <0.1. In the PPI comparison, SMD exceeded 0.1 for 21 covariates after PS stratification, including gastrointestinal hemorrhage, anemia due to blood loss, melena, pregnancy and pregnancy complications, venous varices, diaphragmatic hernia and chronic pulmonary edema. In the hydroxychloroquine comparison, the SMD exceeded 0.1 for the following covariates; age group 30-34, venous varices, distention of vein, ectactic vein, pneumonia and influenza. In the famotidine non-users comparison, all SMDs for baseline characteristics were less than 0.1 after PS stratification. For all baseline characteristics before and after PS adjustment for famotidine users compared to PPI users are plotted in Figure 2. For all baseline characteristics before and after PS adjustment for famotidine users compared to hydroxychloroquine users and to famotidine non-users before and after PS stratification are plotted in Supplement Figures 1, 2 respectively. The complete list of baseline characteristics before and after matching is available in the study link above, under the tab Covariate balance.

**Figure 2:**
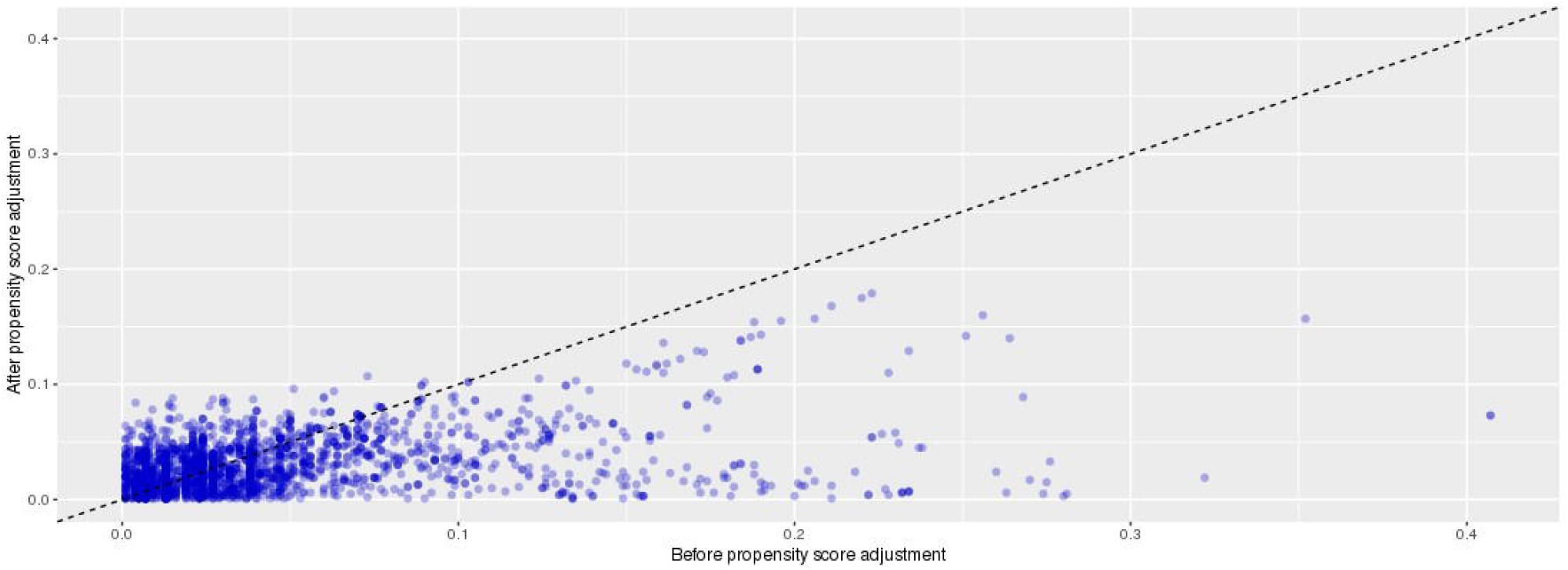
Covariate balance before and after propensity score adjustment for Famotidine users versus proton pump inhibitor (PPI) user. We plotted the absolute standardized difference of population proportions of all available patient characteristics on demographics and conditions before and after propensity score stratification

### Risk of death and death or intensive services

HRs for the relative risk of incidence of death and death or intensive services are presented in Table 3 for the PS stratified and PS matched analyses. The risk of death was not significantly different among famotidine users compared to PPI users with PS stratification (HR 1.14, 95%CI 0.94-1.39). Similarly, the risk of death or intensive services (combined) was similar among the two groups (HR 1.13, 95%CI 0.96-1.32) after PS stratification. When comparing famotidine users to hydroxychloroquine, both the risk of death (HR 1.03, 95% 0.85-1.24) and the risk of death or intensive services (combined) (HR 1.05, 95% 0.90-1.22) were similar between groups after PS stratification. Finally, when comparing famotidine users to non-users, no significant difference in the risk of death (HR 1.03, 95% 0.89-1.18) or the risk of death or intensive services (combined) (HR 1.03, 95% 0.95-1.15) was observed after PS stratification. PS matching HRs followed the same trend.

**Table 3.** Relative risk of death and death or intensive care services for proton pump inhibitor (PPI) users, hydroxychloroquine users, famotidine users and non-users. We report hazard ratios (HRs) and their 95% confidence intervals (CIs) and p-value (P), with propensity score (PS) stratification or matching

|  | PS-stratified |  | PS-matched |  |
| --- | --- | --- | --- | --- |
|  | HR (95% CI) | *P* | HR (95% CI) | *P* |
| ***Famotidine users versus PPI users*** |  |  |  |  |
| **Outcome= death** | 1.14 (0.94 - 1.39) | 0.18 | 1.02 (0.82-1.27) | 0.85 |
| **Outcome= death or intensive services** | 1.13 (0.96 - 1.32) | 0.14 | 1.09 (0.91-1.31) | 0.33 |
| ***Famotidine users versus hydroxychloroquine users*** |  |  |  |  |
| **Outcome= death** | 1.03 (0.85 - 1.24) | 0.79 | 0.98 (0.77-1.23) | 0.85 |
| **Outcome= death or intensive services** | 1.05 (0.90 - 1.22) | 0.55 | 1.12 (0.93-1.37) | 0.24 |
| ***Famotidine users versus non-user*** |  |  |  |  |
| **Outcome= death** | 1.03 (0.89 - 1.18) | 0.67 | 1.03 (0.86-1.24) | 0.74 |
| **Outcome= death or intensive services** | 1.03 (0.92 - 1.15) | 0.62 | 1.00 (0.86-1.16) | 0.99 |

## Discussion

Using real-world data from a large multi-institutional hospital database, we found no evidence of a reduced risk of death among hospitalized COVID-19 patients who used famotidine compared to those who did not or compared to PPI or hydroxychloroquine users. Similarly, there was no observed effect on the composite outcome of death or intensive services when comparing famotidine users to patients in the three comparator groups.

Prior literature on safety and mortality outcomes among COVID-19 patients treated with famotidine is limited to a series of small single-institution studies, and results from these studies are conflicting. Freedberg, et al^6^ found that after PS matching, patients who used famotidine (84 users) were at reduced risk of death or intubation (combined outcome) with a HR of 0.43 (95% CI 0.21-0.88). Another retrospective observational study found that when comparing hospitalized COVID-19 patients who received famotidine (83 patients) to those who did not, famotidine users were at reduced risk of in-hospital mortality (odds ratio OR 0.37, 95% confidence interval 0.16–0.86, P = 0.021) and combined death or intubation (OR 0.47, 95% confidence interval 0.23–0.96, P = 0.040)^7^. However, results from a retrospective cohort study conducted on 51 patients in Hong Kong found no significant association between severe COVID-19 disease and use of famotidine compared to non-users (OR:1.34, 95% CI:0.24– 6.06; p=0.72)^9^. These findings, both for positive and no association, could be potentially attributed to confounding and selection bias in comparator selection, two sources of systematic error that our study sought to address.

We implemented large scale PS adjustment to account for possible measured confounding. We implemented a set of study diagnostics and included 3 different comparators to increase our confidence in the study findings. However, residual confounding due to unobserved factors, such as pre-admission drug use, may still exist, as evidenced by having some SMDs>0.10 even after propensity score adjustment. While PHD contains hospital data from multiple institutions, this study still represents results from one data source and further replication across other sources would increase confidence in the findings. Our study did not consider strength or dose or duration of exposure for any of the exposures and may not generalize to the high dose exposure under investigation in the ongoing clinical trial or in other clinical contexts outside of hospital admission. However, our study findings reflect the real-world use of famotidine in hospitalized COVID-19 patients, and suggests further evidence is needed to demonstrate its real-world effectiveness.

## Data Availability

To protect patient privacy, all patient-level data were maintained securely behind institutional firewalls. Analysis code was executed by researchers, which generated only aggregate summary statistics (cohort counts, model coefficients) which were then synthesized in preparation of this manuscript. All aggregate summary statistics produced have been made publicly available at: https://data.ohdsi.org/Covid19EstimationFamotidine/

## Abbreviations

CDM: Common Data Model
CI: Confidence interval
GERD: Gastroesophageal reflux disease
HRs: hazard ratios
MDRR: minimum detectible risk ratio
OHDSI: Observational Health and Data Sciences and Informatics
OMOP: Observational Medical Outcomes Partnership
PHD: Premier Hospital Database (PHD
PS: propensity score
PPI: proton pump inhibitors
SMD: standardized mean differences

## Figure legends

- *Supplement Figure 1. Covariate balance before and after propensity score adjustment for Famotidine users versus hydroxychloroquine users*. We plotted the absolute standardized difference of population proportions of all available patient characteristics on demographics and conditions before and after propensity score stratification
- *Supplement Figure 2. Covariate balance before and after propensity score adjustment for Famotidine users versus non-users*. We plotted the absolute standardized difference of population proportions of all available patient characteristics on demographics and conditions before and after propensity score stratification

## Acknowledgements

We acknowledge Dr. Akash Pandhare, M.D., Ph.D. for his valuable contribution in summarizing the related medical literature on the COVID-19 immunology.

